# Next weeks of SARS-CoV-2: Projection model to predict time evolution scenarios of accumulated cases in Spain

**DOI:** 10.1101/2020.04.09.20059881

**Authors:** Antonio Monleon-Getino, Jaume Canela-Soler

## Abstract

**Background and objectives:** SARS-CoV-2 is a new type of coronavirus that can affect people and causes respiratory disease, COVID-19. It is affecting the entire planet and we focus in Spain, where the first case was detected at the end of January 2020 and in recent weeks it has increased in many cases. We need predictive models in order to be efficient and take actions. The general goal of this work is present a new model of SARS-CoV-2 to predict different scenarios of accumulated cases in Spain.

**Material and methods:** In this short report is used a model proposed previously, based on a parametric model Weibull and in a the library BDSbiost3 developed in R to infer and predict different scenarios of the evolution of SARS-CoV-2 for the accumulated cases in Spain after the spread that affects Spain detected at the end of January of this year.

**Results:** In the analyses presented, projective curves have been generated for the evolution of accumulated cases in which they reach about 4,000 cases or about 15,000 cases, for which the lines of the day in which the value for 90 will be reached can be seen vertically 90, 95 and 99% of the asymptote (maximum number of cases, from that day they will begin to descend or remain the same), that is why the vertical lines would indicate the brake of the disease. For the worst-case scenario, it takes 118, 126 or 142 days to reach the maximum number of cases (n = 15,000) to reach 90, 95 and 99% of the asymptote (maximum number of cases), respectively. This means translated in a time scale that in the worst case the virus will not stop its progress, in Spain, until summer 2020, hopefully before.

**Comments and conclusions:** This model could be used to plan the resources and see if the policies or means dedicated to the virus are slowing the progress of the virus or it is necessary to implement others that are more effective, and can also validate a method for future outbreaks of diseases such as these.

## 1. Background

SARS-CoV-2 is a new type of coronavirus, a broad family of viruses that normally affect only animals, that can affect people and causes COVID-19. It has been detected for the first time in 2019/12 in the city of Wuhan (China). They produce clinical conditions ranging from the common cold to more serious diseases, such as the coronavirus that caused severe acute respiratory syndrome (SARS-CoV) a few years ago and the coronavirus that causes the Middle East respiratory syndrome (MERS-CoV) [1]. It seems that the transmission would be through contact with infected animals or by close contact with the respiratory secretions that are generated with the cough or sneeze of a sick person.

On January 31, the first confirmed case of SARS-CoV-2 was detected in Spain corresponding to a German citizen who was under observation by the health authorities after learning that he had had close contact with a patient diagnosed with infection by Coronavirus in Germany. These cases have been increasing exponentially, until reaching 999 confirmed cases today [2].

The Spanish Ministry of Health has activated a permanent monitoring commission where the evolution of the coronavirus situation is evaluated. It is in contact with scientific societies for the development of specific protocols in relation to the clinical management of cases. Update daily the report about the outbreak, accessible to the general public [2]. The rate of growth of cases in each country is different. Japan, Hong Kong, Singapore have seen infections grow gradually since January. In other countries such as Spain, France or Germany, cases shot up very quickly following the wake of Italy, which gave the alarm in Europe.

The development of the epidemic follows an exponential growth model in accumulated cases until reaching a maximum point. In China seems that a decline in the most recent days is likely to be due to underascertainment of cases with recent onset and delayed identification and reporting rather than a true turning point in incidence [3]. But can we predict when the maximum number of cases will arrive? Some statements by the Chinese health authorities indicate that the infections will last until summer, but can we apply any model or project these expectations over time?

## 2. Objectives

The general goal of this work is present a new model of SARS-CoV-2 to predict different time scenarios of accumulated cases in Spain.

## 3. Material and methods

In this short report is used a model proposed previously, based on a parametric model Weibull [4, 5, 6] computed with the function Weibull4p.monle1() of the library BDSbiost3[6]. This mathematical function was developed in R to infer and predict different scenarios of the evolution of SARS-CoV-2 for the accumulated cases in Spain after the spread that affects Spain detected at the end of January of this year until last date with information.

This projection offered by function is based on a consistent statistical growth model, used previously in biology [4, 5,6] and can allow authorities and epidemiologists to predict cases and take measures of them, as well as to know the approximate dates for the best and worst scenarios. This method is based on the extrapolation rarefaction curve using a Weibull growth model [4, 5] to estimate the maximum number of accumulated cases of coronavirus as a function of time (days) using numerical methods for non linear models. This approach allows us to compute the effort at different confidence intervals and to obtain an approximate time interval of when the disease will slow its speed in terms of the number of cases accumulated.

This would also be possible for the number of new affected and for cases in which patients recover.

## 4. Results

In the analyzes presented, projective curves (see Figure 1) have been generated for the evolution of accumulated cases in which they reach about 4,000 cases or about 15,000 cases, for which the lines of the day in which the value for 90 will be reached can be seen vertically 90, 95 and 99% of the asymptote (maximum number of cases, from that day they will begin to descend or remain the same), that is why the vertical lines would indicate the slow down of the disease.

**Figure 1:**
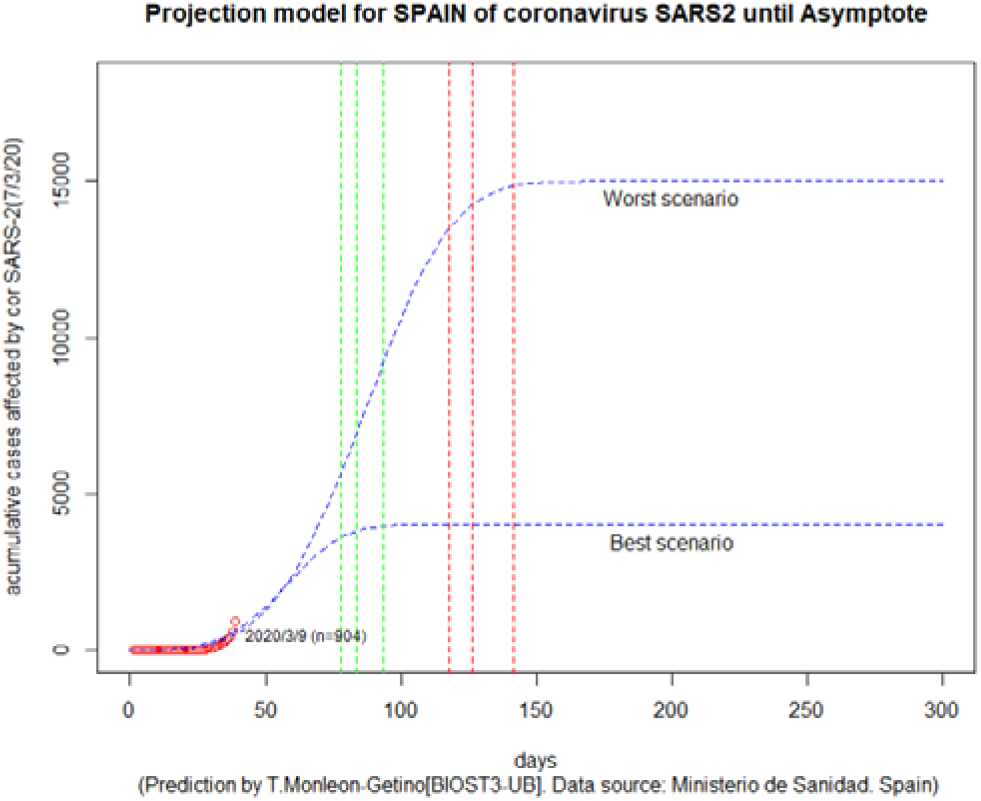
Prediction of accumulated cases of SARS-CoV-2 for the next 300 days based on the use of Weibull model computed with the function Weibull4p.monle1() of the library BDSbiost3 for R. Red dots: real cases. Blue lines: simulation for two scenarios (worst scenario with 15,000 cases, and best scenario for 4,000 cases approx.). Vertical lines indicate the day to reach 90, 95 or 99% asymptote (maximum number of cases). Green vertical line for the best scenario. Red vertical line for the worst scenario.

Thus, for the first scenario (Figure 1), to reach approximately 4,000 cases, the projective model indicates that it will take 78, 84 and 93 days to reach 90, 95 or 99% of the asymptote that in this case would be about 5 times the maximum number of cases (n = 4,000). For the worst-case scenario, it takes 118, 126 or 142 days to reach the maximum number of cases (n = 15,000). This means translated in a time scale that in the worst case the virus will not stop its progress until summer 2020, hopefully before.

## 5. Conclusions and comments

This predictive function could be used to plan the resources and see if the policies or means dedicated to the virus are slowing the progress of the virus or it is necessary to implement others that are more effective, and can also validate a method for future outbreaks of diseases such as these.

The method can easily be validated with the daily updated data for Spain, which can be found at: https://www.epdata.es/datos/coronavirus-china-datos-graficos/498

This method proposed to predict time scenarios and projection could also be used for other countries. The function is freely available to those scientists who want to use it to make their own predictions based on the available data.

## Data Availability

All data are avalaible in https://www.epdata.es/datos/coronavirus-china-datos-graficos/498
http://biost3.blogspot.com/2020/03/modelo-de-prediccion-de-casos-para.html

https://www.epdata.es/datos/coronavirus-china-datos-graficos/498

http://biost3.blogspot.com/2020/03/modelo-de-prediccion-de-casos-para.html

## Notes

### Competing Interest Statement

The authors have declared no competing interest.

### Funding Statement

This work has no financial support of any kind

